# Influence of adherence to social distancing due to the COVID-19 pandemic on physical activity level in post-bariatric patients

**DOI:** 10.1101/2020.08.13.20174458

**Authors:** Diego Augusto Nunes Rezende, Ana Jéssica Pinto, Karla Fabiana Goessler, Carolina Ferreira Nicoletti, Sofia Mendes Sieczkowska, Kamila Meireles, Gabriel Perri Esteves, Rafael Genario, Gersiel Nascimento de Oliveira, Marco Aurélio Santo, Roberto de Cleva, Hamilton Roschel, Bruno Gualano

## Abstract

**Objectives:** To assess physical activity levels in post-bariatric patients who adhered or not to social distancing measures due to the Covid-19 pandemic. Our secondary aim was to compare physical activity estimates between objectively measured and self-reported physical activity level.

**Methods:** In this descriptive, cross-sectional study, we assessed physical activity level using accelerometers and a validated questionnaire in 33 post-bariatric patients who reported to be adherent (n=15) or not (n=18) to social distancing measures.

**Results:** Patients adherent to social distancing measures spent more time in sedentary behavior (1.1 hours/day [95%CI: 0.1, 2.2]; p=0.045) and less time in moderate-to-vigorous physical activity (−12.2 min/day [95%CI: −23.8, −0.6]; p=0.040) compared to non-adherent ones. No difference was observed for light-intensity physical activity. Bland-Altman analysis comparing objective and subjective physical activity estimates showed a bias for time spent in sedentary behavior and moderate-to-vigorous physical activity of 2.8 hours/day and 8.5 min/day, respectively.

**Conclusion:** Post-bariatric patients who were adherent to social distancing measures due to the Covid-19 outbreak were more inactive and sedentary than non-adherent ones. Additionally, questionnaire data widely underestimated sedentary behavior. Strategies to increase or at least sustain physical activity levels in post-bariatric patients exposed to social distancing measures are necessary during the Covid-19 pandemic.

What is already known about this subject?
- Reduced physical activity is associated with poor health-related outcomes in patient undergoing bariatric surgery.
- The impact of the social distancing measures due to Covid-19 pandemic on objectively-measured physical activity in this condition remains unknown.

What are the new findings in your manuscript?
- Adherence to social distancing due to Covid-19 pandemic was associated with decreased objectively measured physical activity and increased sedentary behavior in patients who had undergone bariatric surgery.
- Questionnaire data widely underestimated sedentary behavior when compared to accelerometry data.

How might your results change the direction of research or the focus of clinical practice?
- Given the increased burden of cardiovascular diseases related to inactivity and sedentary behavior, strategies to increase physical activity in post-bariatric patients are clinically relevant during the Covid-19 pandemic.
- The use of validated accelerometers is recommended to screen and track physical activity during the pandemic.

## Introduction

Bariatric surgery is an effective treatment for severe obesity (1), resulting in substantial weight loss, remission of type 2 diabetes (2), and improved overall cardiometabolic profile (3). Weight loss often peaks 1 year following bariatric surgery, and then it is commonly followed by a partial and gradual weight regain after 2 years (3). However, when physical activity is increased during the post-surgery period, patients may exhibit a superior initial weight loss and long-term weight maintenance (4), which is paralleled by sustained improvements in cardiometabolic health (5). Despite the acknowledged benefits of increasing physical activity in the post-surgery period, most patients remain physically inactive and sedentary (6).

As of March 24, 2020, Sao Paulo government adopted a set of social distancing measures (including a “stay-at-home” order) to contain the Covid-19 outbreak. In this scenario, one could expect increases in physical inactivity and sedentary behavior due to social distancing exposure, a condition that could deteriorate the overall health of patients who have undergone bariatric surgery.

To shed light on this, we assessed physical activity levels in post-bariatric patients who reported to be adherent or not to social distancing during the Covid-19 pandemic. As a secondary objective, we compared physical activity estimates between objective and self-reported methods. Our hypotheses were as follows: *(i)* patients who were adherent to social distancing would be more inactive and sedentary compared to those who were non-adherent; *(ii)* self-reported physical activity would show poor agreement with objective estimates (by accelerometer).

## Methods

### Experimental design and participants

We conducted a descriptive, cross-sectional study. Data were collected between April 20 and July 9, 2020, period in which the social distancing measures were in place in Sao Paulo, Brazil. Thirty-three patients who had undergone bariatric surgery were recruited from the outpatient Bariatric Surgery Clinic (Clinical Hospital, School of Medicine, University of Sao Paulo). Inclusion criteria were as follows: being ≥ 18 years old, having a surgery elapsed time ≤ 12 months, and not presenting with any Covid-19 symptoms.

Patients were asked about compliance with social distancing measures by phone interview. They were considered to be “adherent” when they answered *“I always comply with social distancing”* (n= 15), whereas those who answered *“I never comply with social distancing”* or *“I sporadically comply with social distancing”* (n=18) were considered to be “non-adherent”.

This trial was approved by the local ethical committee. Patients signed an informed consent form prior to participation.

### Physical activity level

Physical activity level was measured using ActiGraph GT3X® accelerometers (ActiGraph, Pensacola, Florida) and the International Physical Activity Questionnaire (IPAQ) short version.

All participants were instructed to wear the accelerometer for 7 days during waking hours, except when bathing, on a belt on the right side of the waistline. Data were exported in 60-s epochs using ActiLife 6 software, v. 6.11.9, and analyzed following standard criteria (7). Freedson cut-points were used to classify epochs in sedentary time (<100 counts/min), light-intensity physical activity (≥ 100 and <1952 counts/min), and moderate-to-vigorous physical activity (≥1952 counts/min) (8). Sedentary time was reported as total time per day and also accrued in bouts ≥ 30 min. Data was adjusted to a wear time of 16 hours/day to avoid bias due to differences in wear time among participants.

We administered the IPAQ short version through a phone call interview. The IPAQ short form, which enquires about physical activity and sedentary time in the past seven days. Time spent per day in each activity was calculated as the number of days multiplied by the number of hours reported for each activity and then averaged to seven days. A Brazilian validated version of the questionnaire was used in this study (9).

### Statistical analysis

Dependent variables were tested using independent-sample T-tests. Bland-Altman technique was used to calculate the bias and limits of agreement between accelerometer and IPAQ for sedentary time and moderate-to-vigorous physical activity, using Graph Pad Prism 7. Of note, we decided not to use the Bland-Altman analysis for light-intensity physical activity because IPAQ assesses walking activities exclusively, which certainly underestimates time spent in the light-intensity domain. Data are presented as mean (95% confidence intervals [95%CI]) or absolute and relative frequency (n[%]). Significance level was set at p≤0.05.

## Results

Patients were 48.1 years (44.3, 52.0) and 27 (81.8%) were women. Twenty-five (75.8%) and 8 (24.2%) underwent Y-in-Roux gastric bypass and sleeve gastrectomy surgery, respectively. Post-operative time was 7.1 months (6.2, 8.0) and body mass index was 35.7 kg/m^2^ (33.3, 38.1). Fifteen patients were classified as adherent and 18 as non-adherent to social distancing.

Patients who were adherent to social distancing spent more time in sedentary behavior (1.1 hours/day [95%CI: 0.1, 2.2]; p=0.045) and less time in moderate-to-vigorous physical activity (−12.2 min/day [95%CI: −23.8, −0.6]; p=0.040) compared to non-adherent ones. No difference was observed for prolonged sitting ≥30 min (0.6 hours/day [95%CI: −0.2, 1.5]; p=0.147) and light-intensity physical activity (−0.9 hours/day [95%CI: −1.9, 0.1]; p=0.80). Only three (20%) patients were classified as physically active among those who were adherent to social distancing, whereas 8 (44.4%) were classified as physically active among non-adherent ones.

Grouping all patients, mean time spent in sedentary behavior, light-intensity physical activity, and moderate-to-vigorous physical activity were 9.5 hours/day (9.0, 10.1), 6.1 hours/day (5.6, 6.7), and 19.5 min/day (12.7, 26.3), respectively. Questionnaire data showed that patients spent 4.8 hours/day (3.8, 5.9) in sedentary behavior, 0.2 hours/day (0.1, 0.4) walking, and 8.3 min/day (1.6, 15.0) in moderate-to-vigorous physical activity. Bias for time spent in sedentary behavior and moderate-to-vigorous physical activity were 2.8 hours/day (−2.8, 8.4; Figure 2, panel A) and 8.5 min/day (−28.5, 45.6; Figure 2, panel B), respectively. Bias did not differ between patients who were adherent or non-adherent to social distancing (data not shown).

**Figure 1.**
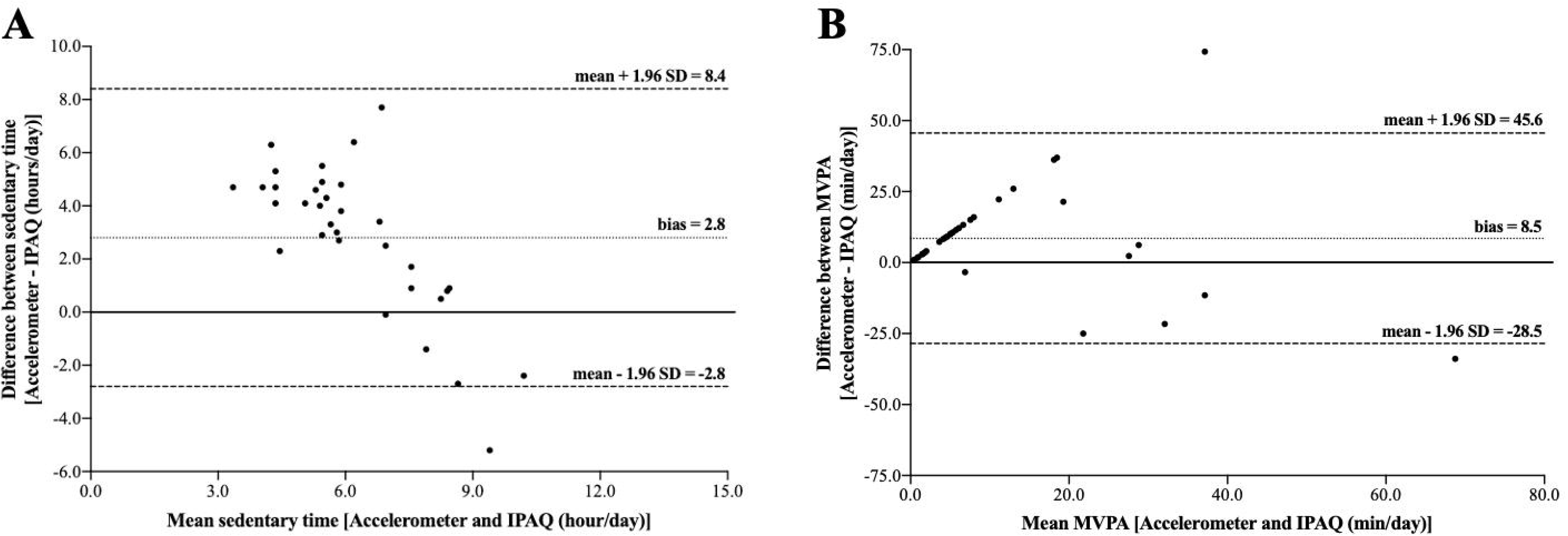
Time spent in sedentary behavior (panel A), light-intensity physical activity (panel B) and moderate-to-vigorous physical activity in post-bariatric patients who were adherent and non-adherent to social distancing measures.

**Figure 2.**
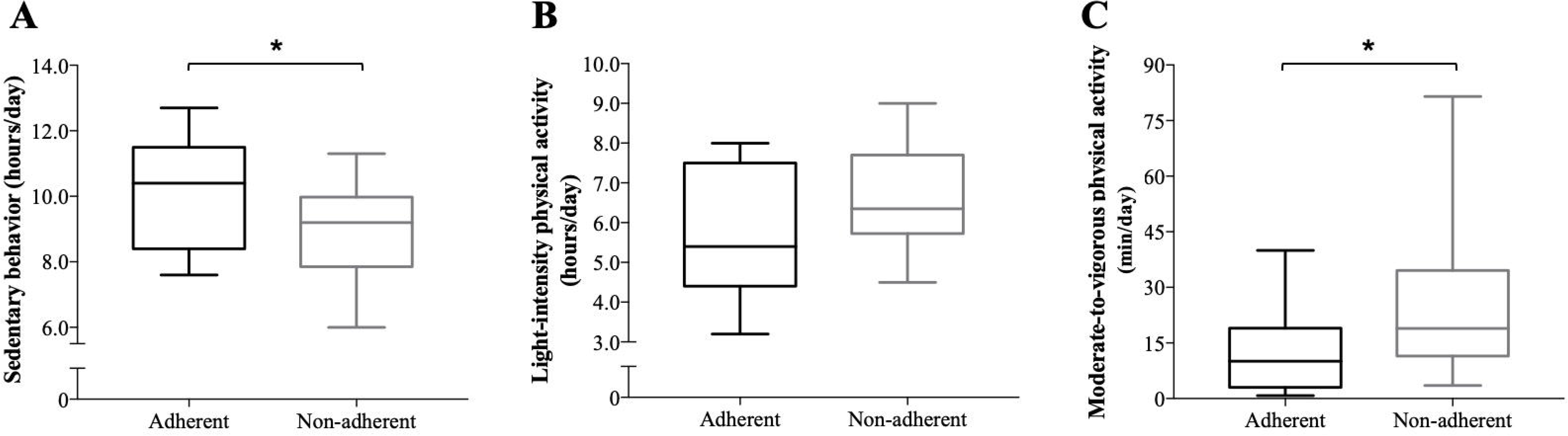
Bland-Altman plot for time spent in sedentary behavior (panel A) and moderate-to-vigorous physical activity (panel B). Abbreviation: IPAQ, International Physical Activity Questionnaire; MVPA, moderate-to-vigorous physical activity.

## Discussion

To our knowledge, this is the first study to assess physical activity levels using validated accelerometers in post-bariatric patients during the Covid-19 pandemic. Our main findings suggest that post-bariatric patients who were adherent to social distancing were more inactive and sedentary than those who were non-adherent. Physical inactivity and sedentary behavior are potential risk factors that could deteriorate cardiometabolic health in post-bariatric patients and is of particular importance to those exposed to social distancing measures during the Covid-19 pandemic.

The data showing that patients exposed to social distancing spent less time in moderate-to-vigorous physical activity and more time in sedentary behavior suggest that the set of social distancing measures to prevent the spread of Covid-19 has come at detriment of physical activity. Inactivity and sedentary behavior associate with chronic diseases and all-cause mortality (10, 11). Even short-term exposure (14 days) to inactivity and sedentariness results in metabolic dysfunction, muscle wasting, fat accumulation, and impaired physical capacity in healthy individuals (12). In patients who have undergone bariatric surgery, the repercussion of physical inactivity and sedentary behavior during the post-operative period is underexplored; however, higher physical activity has been associated with better cardiometabolic risk factors (5). Furthermore, exercise training has been shown to improve insulin resistance (13), vascular function (14), and inflammatory markers (14), and attenuate the loss of bone and muscle mass (15, 16). Thus, increasing physical activity could be a key strategy to improve overall health among post-bariatric patients and are confined during the pandemic.

Physical activity questionnaires are commonly used due to their easy-of-use and low cost. However, questionnaires might be misleading due to their poor validity and reliability (17). When compared to objectively measured physical activity, individuals usually overreport their time spent in physical activity (18), whereas underreport sedentary time (19), which seems to be more pronounced among individuals with obesity (18). To our knowledge, our study was the first to compare sedentary behavior data between objective and self-reported tools in post-bariatric patients. Patients underreported their sedentary time by 2.8 hours/day (29.5%), concurring with data from a recent meta-analysis (19). Interestingly, however, our patients underreported moderate-to-vigorous physical activity by 8.5 min/day (42.9%), contrasting with previous literature (18, 20). Due to the expected difficulty of collecting objective physical activity data during the Covid-19 pandemic, researchers might be inclined to use questionnaires. However, healthcare professionals and police-makers should exercise caution when interpreting these data as the agreement of self-reported and objective methods is widely poor, as evidenced by the current study.

The main strength of this study is the concomitant use of validated accelerometer and questionnaire, which allowed characterizing physical activity as well as identifying the limitation of assessing physical activity using subjective methods during the Covid-19 pandemic. Also, the comparison between patients who were adherent and non-adherent to social measures enhances our ability to establish causation. The limitations of this study include the reduced sample size and the cross-sectional nature of the study, which hampers a definitive conclusion on the causal effect of social distancing on physical activity.

In conclusion, post-bariatric patients who were adherent to social distancing to contain the Covid-19 pandemic were more inactive and sedentary than those who were non-adherent. Given the increased burden of cardiovascular diseases related to inactivity and sedentary behavior, strategies to increase physical activity level in post-bariatric patients, particularly those exposed to social distancing, are of emergent need during the Covid-19 pandemic.

## Data Availability

The datasets used and/or analyzed during the current study will be available from the corresponding author on reasonable request.

## Acknowledgements

A.J.P., K.F.G., S.M.S., and B.G. were supported by São Paulo Research Foundation – FAPESP (grants #2015/26937-4, #2019/18039-7, #2019/15231-4, #2017/13552-2). C.F.N. was supported by National Council for Scientific and Technological Development – CNPq (grant #402123/2020-4). D.R. and. K.M. were supported by Coordenação de Aperfeiçoamento de Pessoal de Nível Superior – CAPES – Finance Code 001.

